# Colorectal cancer risk stratification using a polygenic risk score in symptomatic patients presenting to primary care – a UK Biobank retrospective cohort study

**DOI:** 10.1101/2023.12.08.23299717

**Authors:** Bethan Mallabar-Rimmer, Samuel WD Merriel, Amy P Webster, Andrew R Wood, Matthew Barclay, Jessica Tyrrell, Katherine S Ruth, Christina Thirlwell, Richard Oram, Michael N Weedon, Sarah ER Bailey, Harry D Green

## Abstract

Colorectal cancer (CRC) is a leading cause of cancer mortality worldwide. Accurate cancer risk stratification approaches could increase rates of early CRC diagnosis, improve health outcomes for patients and reduce pressure on diagnostic services. The faecal immunochemical test (FIT) for blood in stool is widely used in primary care to identify symptomatic patients with likely CRC. However, there is a 6–16% noncompliance rate with FIT in clinic and ∼90% of patients over the symptomatic 10µg/g test threshold do not have CRC.

A polygenic risk score (PRS) quantifies an individual’s genetic risk of a condition based on many common variants. Existing PRS for CRC have so far been used to stratify asymptomatic populations. We conducted a retrospective cohort study of 53,112 UK Biobank participants with a CRC symptom in their primary care record at age 40+. A PRS based on 207 variants, 5 genetic principal components and 24 other risk factors and markers for CRC were assessed for association with CRC diagnosis within two years of first symptom presentation using logistic regression. Associated variables were included in an integrated risk model and tested for ability to predict CRC diagnosis within two years, using receiver operating characteristic area under the curve (ROCAUC) and Akaike information criterion (AIC).

An integrated risk model combining PRS, age, sex and patient-reported symptoms was highly predictive of CRC development (ROCAUC: 0.80, 95% confidence interval: 0.78– 0.81). This model has the potential to improve early diagnosis of CRC, particularly in cases of patient non-compliance with FIT.

**Lay Abstract:** Bowel cancer is one of the most common types of cancer worldwide, and patients diagnosed earlier have a much better chance of survival. Finding ways to predict which people are at risk of developing bowel cancer is therefore a research priority.

In this study, we used genetics and information about patients (such as age and sex) to predict which patients are at high risk of developing bowel cancer within two years of seeing their GP with a symptom. We tested 30 risk factors and identified eight that were more common in patients who developed bowel cancer shortly after experiencing symptoms.

These eight risk factors included: older age, being male, larger waist circumference, smoking, higher inherited genetic risk, and presence of two symptoms – change in bowel habit (including constipation or diarrhoea) and/or bleeding from the rectum. On the other hand, stomach pain was the symptom which occurred least in people who developed bowel cancer.

Six of the above risk factors, when combined into one measure of risk (called ‘a risk model’) were good at predicting which patients would develop bowel cancer shortly after symptoms. These factors included age, sex, genetic risk, bleeding from the rectum, change in bowel habit and stomach pain.

This risk model could help doctors decide which symptomatic patients to send for bowel cancer testing. This would allow earlier detection of bowel cancer which would improve outcomes for patients.

## Introduction

Colorectal cancer (CRC) is the second most common cause of cancer mortality in the UK and worldwide.(1,2) In the UK, only 37–41% of CRC cases are diagnosed early, at stage 1 or 2.(3) Earlier diagnosis improves prognosis, and is a research priority for CRC(4,5) and an NHS target for all cancers.(6) Since 67% of UK CRC cases are diagnosed through primary care,(7) early diagnosis in this setting has potential to improve patient outcomes.

### Current diagnostic practice

Patients referred urgently for CRC diagnosis often receive a computed tomography scan or endoscopy procedure (e.g., colonoscopy, flexible sigmoidoscopy).(8) One study of an NHS Trust between 2009 and 2014 showed that <10% of patients referred urgently for suspected CRC received a CRC diagnosis.(9) In 2021, 540,867 colonoscopies were performed in the UK, of which 90.88% were for diagnostic or therapeutic purposes (the remaining 9.12% were for screening).(10) Unnecessary colonoscopies have high cost(11), detrimental environmental impact(12), and may cause patients avoidable discomfort and distress.(13) There is a clear need for improved targeting of CRC diagnostic procedures to patients most at risk. CRC risk stratification could improve early diagnosis rates and minimise unnecessary procedures, thereby reducing burden on patients, clinicians and healthcare providers.(14)

National Institute for Health and Care Excellence (NICE) guidelines, published in August 2023, recommend stratifying primary care patients with bowel symptoms for CRC diagnosis using the quantitative faecal immunochemical test (FIT) for blood in stool.(15) NICE estimate this will prevent 94,291 colonoscopies per year.(16) FIT is offered based on symptoms which vary by age (abdominal mass, change in bowel habit, or iron-deficiency anaemia for patients of all ages; abdominal pain and unexplained weight loss if age 40+; rectal bleeding and one of abdominal pain or weight loss if 50+; anaemia if 60+).(15) Extensive evidence supports FIT as highly sensitive and specific for CRC risk.(17–19) However, FIT uptake varies by age, sex, ethnicity and socioeconomic status, with a 6.4–16.2% noncompliance rate in-clinic.(15,20) Additionally, an evaluation of FIT in clinic showed that of 618 patients with a FIT result over the symptomatic 10ug/g threshold, only 43 (6.97%) had CRC.(21) This study presents a novel method of patient risk stratification in primary care using genetics, symptoms, and patient characteristics, with potential to complement existing methods such as FIT in cases of noncompliance or uncertain results.

### Genetic and integrated risk models to stratify patient risk of CRC

Risk models including both genetic and environmental risk factors have more power to discriminate between CRC cases and controls, compared to models only including environmental risk factors.(22)

A polygenic risk score (PRS) quantifies an individual’s genetic risk of a condition based on many variants.(23) To date, PRS for CRC have screened for CRC risk in asymptomatic populations.(24–27) CRC symptoms are often non-specific for cancer – e.g., abdominal pain, weight loss – and can make it challenging for clinicians to decide which patients may benefit from a diagnostic referral.(28) It has previously been shown that a PRS can stratify symptomatic patients according to risk of developing prostate cancer within a two year window.(29) Therefore, in this study, an integrated risk model (IRM) was developed which combines information about participant symptoms with a PRS and other risk factors, to predict which of a cohort of symptomatic patients are most at risk of CRC in the next two years.

### Aim

The aim of this study was to build an IRM – including environmental risk factors, patient demographics, symptoms, and a PRS – to predict which primary care patients with CRC symptoms will be diagnosed with CRC within two years of their first symptom. The intended use of the IRM is to enable clinicians to decide which symptomatic patients to refer for CRC testing, improving triage of patients in primary care.

### Reporting standards

This report has been written in line with the Polygenic Risk Score Reporting Standards published by the Polygenic Score Catalogue and the Clinical Genome Resource Complex Disease Working Group.(30)

## Subjects and Methods

### Study design and cohort

This was a retrospective cohort study using primary data from UK Biobank (UKBB). UKBB is a database of 500,000 individuals recruited at ages 40–69, between 2006 and 2010, described extensively elsewhere.(31) General practice (GP) records between 1938 and 31 August 2017 were available for ∼230,000 participants.(32,33)

All analysis in this study was completed on the UKBB Research Analysis Platform on DNAnexus, using R coding language version 4.1.1. The study cohort included UKBB participants with at least one CRC symptom in their GP record at or after the age of 40. Symptomatic participants were identified by searching UKBB GP records for Read v2 and v3 codes(34) for the following CRC symptoms: abdominal mass, abdominal pain, abnormal faecal occult blood (FOB) test result, appetite loss, change in bowel habit, iron deficiency, low haemoglobin, rectal bleeding, weight loss. Low haemoglobin was defined as <11 grams per decilitre (g/dl) in females and <13g/dl in males (using self-reported sex of patients).(35) FOB is a test for blood in stool, and was the precursor to FIT.(36) FIT results are unavailable in the UKBB GP records as FIT was introduced in 2017 (the same year UKBB GP records end). The number of Read codes for each CRC symptom, and the number of participants in the final study cohort with each symptom, is listed in Supplementary Table 1. For the full list of Read codes, see the ‘Data Availability’ section of this manuscript.

‘Index date’ refers to the date of a participant’s first recorded CRC symptom, at or after age 40. The age threshold of 40 when searching participants’ symptom records was used because very few UKBB participants were diagnosed with CRC younger than this (Supplementary Figure 1). The cohort was divided into cases, who had a CRC diagnosis within two years of index date, or controls, with no CRC in that period. A CRC diagnosis was defined by searching for ICD-10 codes in participants’ Cancer Registry, Hospital Inpatient Data, and Death records, or for Read codes describing CRC in GP records (see Data Availability). The ICD-10 codes used referred to malignancies of the colon (C18), rectosigmoid junction (C19), rectum (C20) and anus or anal canal (C21). Participants were excluded from the study if they had CRC before the index date, or if they died of a cause other than CRC within two years, as it cannot be determined whether they would have developed CRC. Study design is summarised with a flowchart in Figure 1.

**Figure 1.**
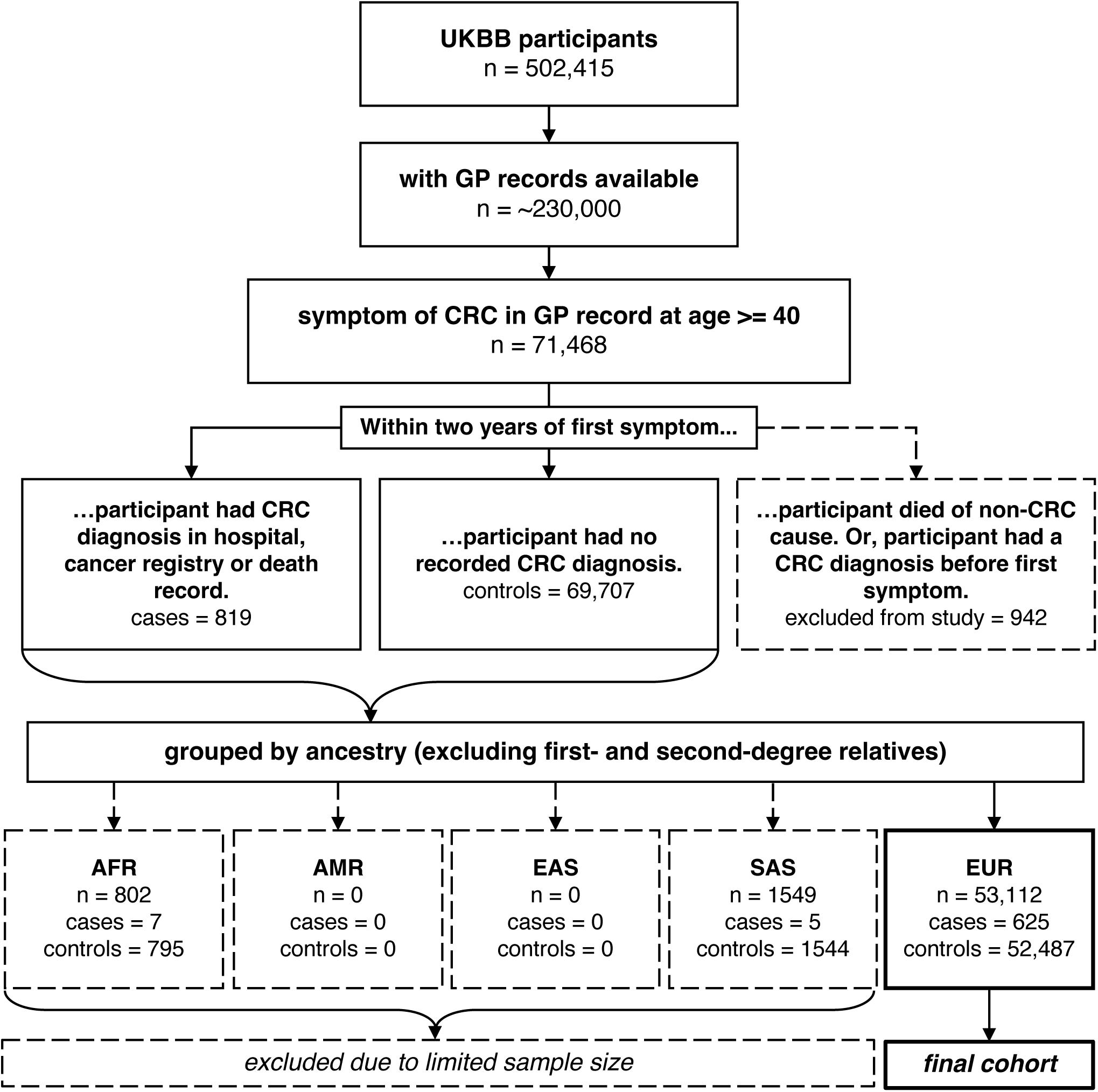
Flowchart of study design. Participants with a CRC symptom in their GP record at age 40+ were included in the study. Cases had a CRC diagnosis within two years of first symptom, whereas controls did not. Excluded participants: died within two years of first symptom (not from CRC), had CRC before first symptom, had non-European ancestry (excluded due to limited case numbers), or were related to the first- or second-degree. Abbreviations: AFR = African, AMR = Admixed America, CRC = colorectal cancer, EAS = East Asian, EUR = European, GP = general practice, SAS = South Asian, UKBB = UK Biobank.

### Ancestry

UKBB participants with European (n=379,768), South Asian (n=9,455), African (n=7,440), East Asian (n=2,481), and Admixed American (n=467) ancestry, excluding individuals related to the first or second degree, were determined using principal components analysis and KING kinship.(37) Following filtering for availability of GP records and presence of a CRC symptom, the European cohort included 625 cases and 52,487 controls, the African cohort 7 cases and 795 controls, the South Asian cohort 5 cases and 1544 controls, and there were no cases or controls with either East Asian or Admixed American ancestry. Due to this limited sample size, the PRS and IRM could only be developed and assessed for predictivity in the European cohort. However, PRS distribution in UKBB was assessed across all five ancestral superpopulations (see Results).

### Participant demographics

The final cohort consisted of 53,112 participants (625 cases and 52,487 controls) who were related to no more than the third degree, had European ancestry, and had a CRC symptom recorded in their GP record between the ages of 40–79 (no UKBB participants had a CRC symptom recorded at 80 or older). The cohort included male and female participants. Incidence of CRC in the cohort (percentage of cases) was 1.18% (0.59% per year). This is higher than annual CRC incidence in the UK (0.13% in 2017, non-age standardised),(38) likely because this study assesses a middle-aged, symptomatic population.

Further sensitivity analyses were conducted by age at first symptom (stratified by decades 40–49, 50–59, 60–69, and 70–79) and sex. Overall, the ability of the PRS and IRM to predict CRC diagnosis within two years of first symptom was tested in a total of 14 subcohorts (4 grouped by age, 2 by sex, and 8 by both age and sex) as well as the full cohort. Age and sex distributions are summarised in Table 1 for the full cohort (Supplementary Table 2 for subcohorts).

**Table 1.** age, sex, and numerical variable distribution of cases and controls in the full cohort. Distributions of these variables are reported for the full cohort and all subcohorts in Supplementary Table 2. Categorical variable distribution across all cohorts is reported in Supplementary Table 5. Abbreviations: N = number of participants, SD = standard deviation.

|  | Cases | Controls | Full Cohort |
| --- | --- | --- | --- |
| **N (% of cohort)** | 625 (1.18%) | 52,487 (98.82%) | 53,112 (100%) |
| **Sex (patient self-reported)** |  |  |  |
| Female | 257 (41.12%) | 31,335 (59.70%) | 31,592 (59.48%) |
| Male | 368 (58.88%) | 21,154 (40.30%) | 21,522 (40.52%) |
| **Age at UKBB baseline/recruitment (years)** |  |  |  |
| Mean (+/- SD) | 50.09 (+/- 7.01) | 51.78 (+/- 7.25) | 51.78 (+/- 7.25) |
| **Age at first CRC symptom (years)** |  |  |  |
| Mean (+/- SD) | 61.09 (+/- 7.76) | 54.91 (+/- 8.77) | 54.98 (+/- 8.79) |
| **Body mass index (BMI) (kg/m<sup>2</sup>)** |  |  |  |
| Mean (+/- SD) | 27.91 (+/- 4.91) | 27.75 (+/- 4.98) | 27.76 (+/- 4.98) |
| **Waist circumference (cm)** |  |  |  |
| Mean (+/- SD) | 93.68 (+/- 14.24) | 90.61 (+/- 13.76) | 90.65 (+/- 13.77) |
| **Townsend deprivation index (TDI)** |  |  |  |
| Mean (+/- SD) | -1.75 (+/- 2.91) | -1.46 (+/- 2.95) | -1.47 (+/- 2.94) |

### Genetic data

UKBB participants were genotyped at ∼850,000 variants with the UKBB Axiom Array, and a further ∼96 million variants were imputed (steps detailed by Bycroft et al.)(31) PRS construction in this study used participant genotypes imputed by the Wellcome Trust Centre for Human Genetics, version 3 in UKBB.(39)

### Polygenic risk score construction

A PRS quantifying participants’ genetic risk of CRC was calculated using 207 genetic variants associated with long-term risk of developing CRC, and their betas (the log odds ratio of the association between variant and phenotype), published by Fernandez-Rozadilla et al. in recent genome-wide meta-analysis.(40) The meta-analysis included 100,204 cases with a diagnosis of CRC, and 154,587 controls without, from ∼90 studies. Inclusion criteria varied by study. 73% of individuals in the meta-analysis were European and 27% were East Asian, mostly matching the European ancestry of our cohort. 95.2% of cases and 86.9% of controls were not from UKBB, reasonably avoiding overfitting with UKBB data.

For each participant in UKBB, a PRS was calculated by scoring dosage (the expected/predicted genotype following imputation) at each of the 207 genetic variants, multiplying each score by the beta for the variant, and summing these values.

### Non-genetic variables and logistic regression analysis

A total of 30 variables (the PRS and 29 others derived from UKBB data-fields) were tested individually for association with CRC diagnosis within the two-year period, using logistic regression. Variables included participant characteristics (age at first CRC symptom and sex), lifestyle variables (Townsend deprivation index, smoking, alcohol intake, and processed meat intake), health measures (body mass index, waist circumference, and whether the participant self-reported having diabetes), PRS, which symptom/s the participant reported at index date, and whether the participant reported a history of bowel cancer in either parent. The first five genetic principal components in UKBB were also included as variables, to test for confounding. All variables were measured at baseline (UKBB recruitment), except genetic variables (genotyping data was released in 2017, imputed genotyping data in 2022 – Supplementary Table 3) and symptoms at index/age at first symptom. All variables had values for >99% of participants (Supplementary Table 4) – any missing values were omitted from logistic regression analysis.

We tested association in the full cohort and 14 subcohorts. Variables significantly associated with either the case or control group (p-value <1.7e-03, Bonferroni corrected threshold) were further tested for ability to predict CRC diagnosis when included in an IRM.

### IRM description and fitting

The IRM estimates the cross-sectional risk of a patient with CRC symptoms either being diagnosed with the cancer within two years of first symptom or not. The IRM was developed for optimal performance according to two measures; receiver operating characteristic area under the curve (ROCAUC) and Akaike information criterion (AIC).

The IRM was constructed by starting with the variable with highest ROCAUC and iteratively adding variables in order of which cause the largest increase in ROCAUC. A second method of IRM construction involved building all possible risk models and ranking these according to AIC score – where a lower AIC score indicates a model which better fits the dataset while excluding inessential variables. Using AIC to penalise models including more variables reduces overfitting.

## Results

### Symptoms and factors associated with CRC risk

In the full cohort, nine variables were associated (p<1.7e-03, Bonferroni-corrected threshold) with increased risk of CRC diagnosis within two years of first symptom presentation, relative to other symptomatic patients in the cohort. These included: older age at index date; higher PRS; self-reported sex being male; increased waist circumference; having ever smoked; smoking previously (as opposed to never having smoked or being a current smoker); an abnormal FOB test result; rectal bleeding; and change in bowel habit. The finding that previous smokers had increased risk of CRC within the two-year period compared to those who reported being current smokers may indicate underlying bias or confounding in this variable; e.g. previous smokers may have smoked more heavily or for more years on average. This variable was therefore excluded from the IRM. FOB test result was also excluded, because this study aimed to assess the IRM as a standalone risk prediction tool, not relying on existing diagnostic tests

One variable, abdominal pain, was more prevalent in controls than cases, and was therefore associated (p=2.10e-69) with decreased risk of CRC diagnosis within two years relative to participants reporting other symptoms. CRC incidence in participants with abdominal pain was 0.21% (∼0.11% per year) – Supplementary Table 5 – similar to annual CRC incidence in the UK of 0.13%.(38) Table 2 shows the p-values, odds ratios, individual ROCAUC scores, and 95% confidence intervals of the ten variables associated with either the case or control group following logistic regression. Supplementary Table 6 shows the results of logistic regression for all 30 variables in the full cohort and subcohorts.

**Table 2:** p-values, odds ratios, ROCAUC, and 95% confidence intervals for all variables shown via logistic regression to be associated with the case or control group, in the full cohort. For logistic regression results in subcohorts, see Supplementary Table 6. 95% confidence intervals are shown in brackets. Risk factors and markers in the table are ordered according to strength of p-value. A p-value threshold of <1.7e-03 follows Bonferroni correction of α = 0.05 for 30 tests. Abdominal pain has an odds ratio <1, showing that participants with abdominal pain had decreased risk of CRC within two years of first symptom relative to the rest of the cohort. Additionally, results suggest that current smokers had decreased risk of CRC diagnosis within two years of first symptom relative to participants in the cohort who self-identified as previous smokers. This may be due to confounding; for example, it is possible that previous smokers may have smoked more heavily or for longer on average than participants who reported currently smoking. Abbreviations: CRC = colorectal cancer, PRS = polygenic risk score, ROCAUC = receiver operating characteristic area under the curve.

| **Risk factor/marker** | **p-value** | **Odds Ratio** | **ROCAUC** |
| --- | --- | --- | --- |
| **Faecal occult blood test abnormal** | $1.6 \times 10^{-132}$ | 12.34 (10.09-15.08)<br>*if symptom recorded at index date* | 0.59 (0.58-0.61) |
| **Abdominal pain** | $2.1 \times 10^{-69}$ | 0.19 (0.15-0.22)<br>*if symptom recorded at index date* | 0.69 (0.68-0.71) |
| **Age at first CRC symptom** | $8.6 \times 10^{-64}$ | 1.09 (1.08-1.10)<br>*per year increase* | 0.70 (0.68-0.72) |
| **PRS** | $1.3 \times 10^{-28}$ | 2.15 (1.88-2.46)<br>*per quintile increase* | 0.63 (0.60-0.65) |
| **Rectal bleeding** | $1.1 \times 10^{-27}$ | 2.51 (2.13-2.96)<br>*if symptom recorded at index date* | 0.59 (0.57-0.61) |
| **Sex** | $3.7 \times 10^{-20}$ | 2.12 (1.81-2.49)<br>*if male* | 0.59 (0.57-0.61) |
| **Waist circumference** | $3.0 \times 10^{-8}$ | 1.02 (1.01-1.02)<br>*per cm increase* | 0.56 (0.54-0.59) |
| **Smoking (current, previous, or never)** |  |  |  |
| never -> previous | $5.0 \times 10^{-6}$ | 1.47 (1.25-1.74)<br>*if previous smoker* | 0.55 (0.53-0.57) |
| never -> current | $8.90 \times 10^{-1}$ | 1.02 (0.77-1.36)<br>*if current smoker* | 0.5 (0.48-0.52) |
| previous -> current | $1.20 \times 10^{-02}$ | 0.69 (0.52-0.92)<br>*if current smoker* | 0.53 (0.51-0.55) |
| **Change in bowel habit** | $7.5 \times 10^{-6}$ | 1.70 (1.35-2.14)<br>*if symptom recorded at index date* | 0.53 (0.51-0.54) |
| **Smoking (ever or never)** | $1.2 \times 10^{-3}$ | 1.32 (1.12-1.56)<br>*if ever smoked* | 0.53 (0.51-0.55) |

### IRM evaluation

The ten aforementioned variables (Table 2), minus smoking status and FOB test result, were used to construct an IRM. After applying AIC scoring to all 255 possible combinations of eight variables, an IRM combining six variables – age at first symptom, sex, PRS, and whether or not the patient reported symptoms of abdominal pain, rectal bleeding, or change in bowel habit – had the lowest AIC score (Table 3). This IRM had a ROCAUC of 0.80, with a 95% confidence interval (CI95) of 0.78–0.81. This was the joint-highest ROCAUC of all 255 models, showing good agreement between ROCAUC and AIC.

**Table 3:** The five IRMs with lowest AIC scores in the full cohort. 255 IRMs were constructed using all possible combinations of the following eight variables: age at first symptom, sex, PRS, smoking (ever or never), waist circumference, and whether the patient reported symptoms of abdominal pain, change in bowel habit, and/or rectal bleeding at index date. AIC scoring was applied to all 255 models. Models which better fit the dataset, while excluding extraneous variables, have lower AIC scores. An individual AIC score cannot be meaningfully interpreted without comparison to other AIC scores. AIC scores across all 255 models ranged from -86639.52 to -85176.01 (range = 1463.51). The five IRMs with lowest AIC scores in each subcohort are shown in Supplementary Table 7. Variables significantly associated (logistic regression p<1.7e-03, Bonferroni-corrected threshold) with either the case or control group vary by subcohort. Only variables with significant association were used to construct IRMs. Therefore, the IRMs tested using AIC scoring are different in each subcohort. Abbreviations: AIC = Akaike information criterion, CI95 = 95% confidence interval, IRM = integrated risk model, PRS = polygenic risk score, ROCAUC = receiver operating characteristic area under the curve.

| IRM (number of variables) | AIC score | ROCAUC (CI95) |
| --- | --- | --- |
| age, abdominal pain, PRS, sex, rectal bleeding, change in bowel habit (6) | -86639.52 | 0.78 (0.80-0.81) |
| age, abdominal pain, PRS, sex, change in bowel habit (5) | -86637.96 | 0.78 (0.80-0.81) |
| age, abdominal pain, PRS, sex (4) | -86628.36 | 0.78 (0.80-0.81) |
| age, abdominal pain, PRS, sex, rectal bleeding (5) | -86626.37 | 0.78 (0.80-0.81) |
| age, abdominal pain, PRS, change in bowel habit (4) | -86600.68 | 0.77 (0.79-0.81) |

The other method of IRM construction used in this study involved adding variables to the IRM in order of which caused the largest increase in ROCAUC. Figure 2 shows that, in the full cohort, an IRM including only four variables (age at first symptom, sex, PRS, and whether or not the patient reported abdominal pain) reaches a ROCAUC of 0.80 (CI95=0.78–0.81), equivalent to the aforementioned six-variable IRM.

**Figure 2.**
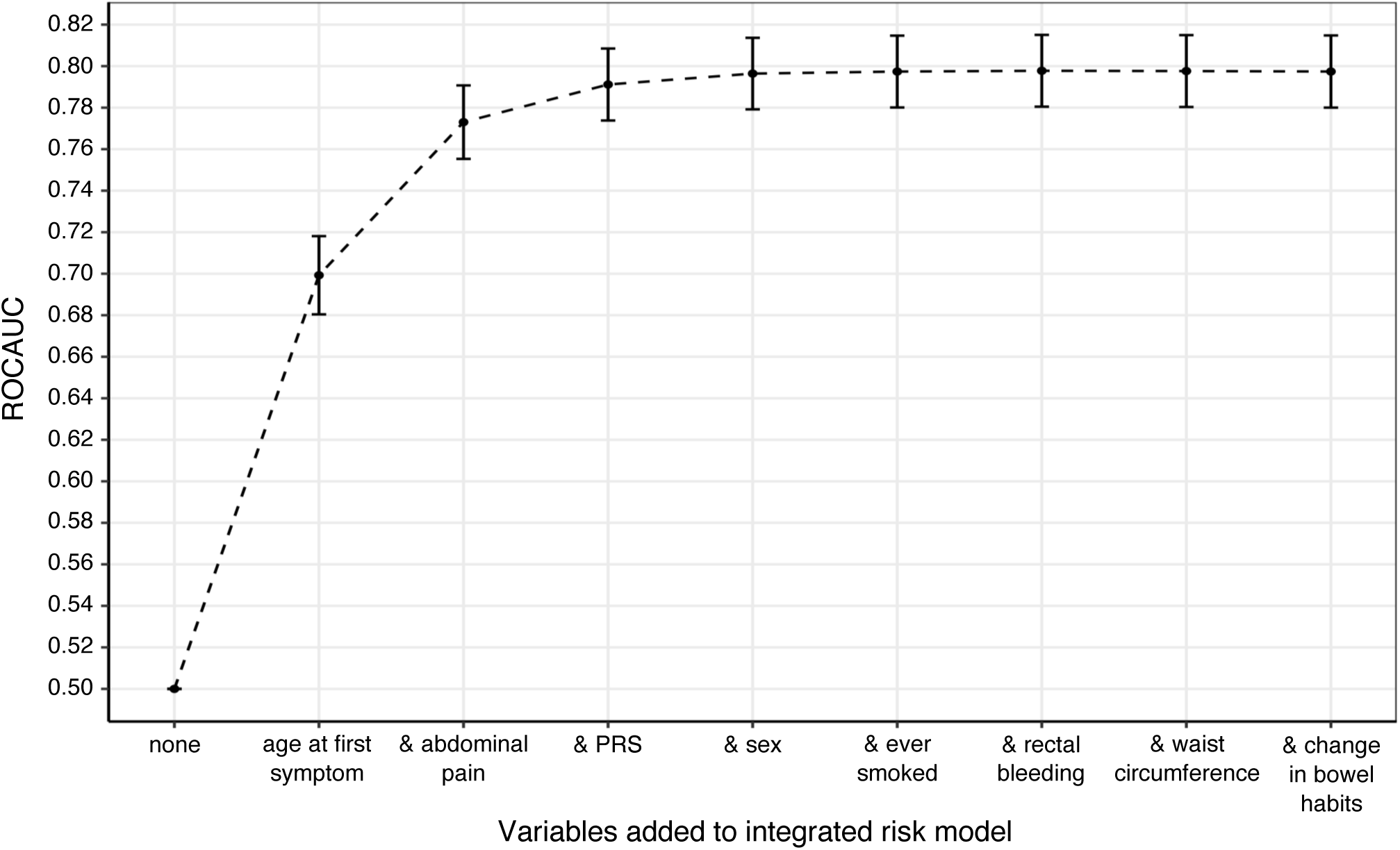
As variables are added to the IRM, ROCAUC increases until plateauing after the inclusion of four variables. The four most predictive variables (age at first symptom, abdominal pain, PRS and sex) had combined ROCAUC of 0.80 (CI95: 0.78-0.81). Abbreviations: PRS = polygenic risk score, ROCAUC = receiver operating characteristic area under the curve.

However, the six-variable IRM had higher ROCAUC across subcohorts (mean: 0.75, standard deviation (SD): 0.04) than the four-variable IRM (mean: 0.74, SD: 0.04). The six-variable IRM had reduced predictivity and wider confidence intervals in participants aged 40-49 than other age groups (Supplementary Figure 2). When splitting the full cohort by sex but not age, ROCAUC was equivalent in female and male participants (0.78, CI95=0.76–0.81). Supplementary Table 7 shows results of IRM evaluation across cohorts.

### PRS evaluation

The PRS had a ROCAUC of 0.63 (CI95=0.60–0.65) in the full cohort, showing moderate ability to discriminate between symptomatic participants with or without CRC. PRS distribution was higher in cases than controls (OR and CI95 >1) in the full cohort and all subcohorts except female participants aged 40–49 (Supplementary Table 6 and Supplementary Figure 3). Logistic regression showed weak statistical evidence for an effect in male participants aged 40-49 (p=9.3e-03) and female participants aged 70-79 (p=1.4e-02), and no evidence of an effect in female participants aged 40-49 (p=9.4e-01). In every other subcohort, p-values met the Bonferroni-corrected value of <1.7e-03 (Supplementary Table 6).

Figure 3 shows that in the full cohort, 1.95% of participants with a PRS in the highest quintile were diagnosed with CRC within two years of first reported symptom, compared to 0.61% of participants with a PRS in the lowest quintile.

**Figure 3.**
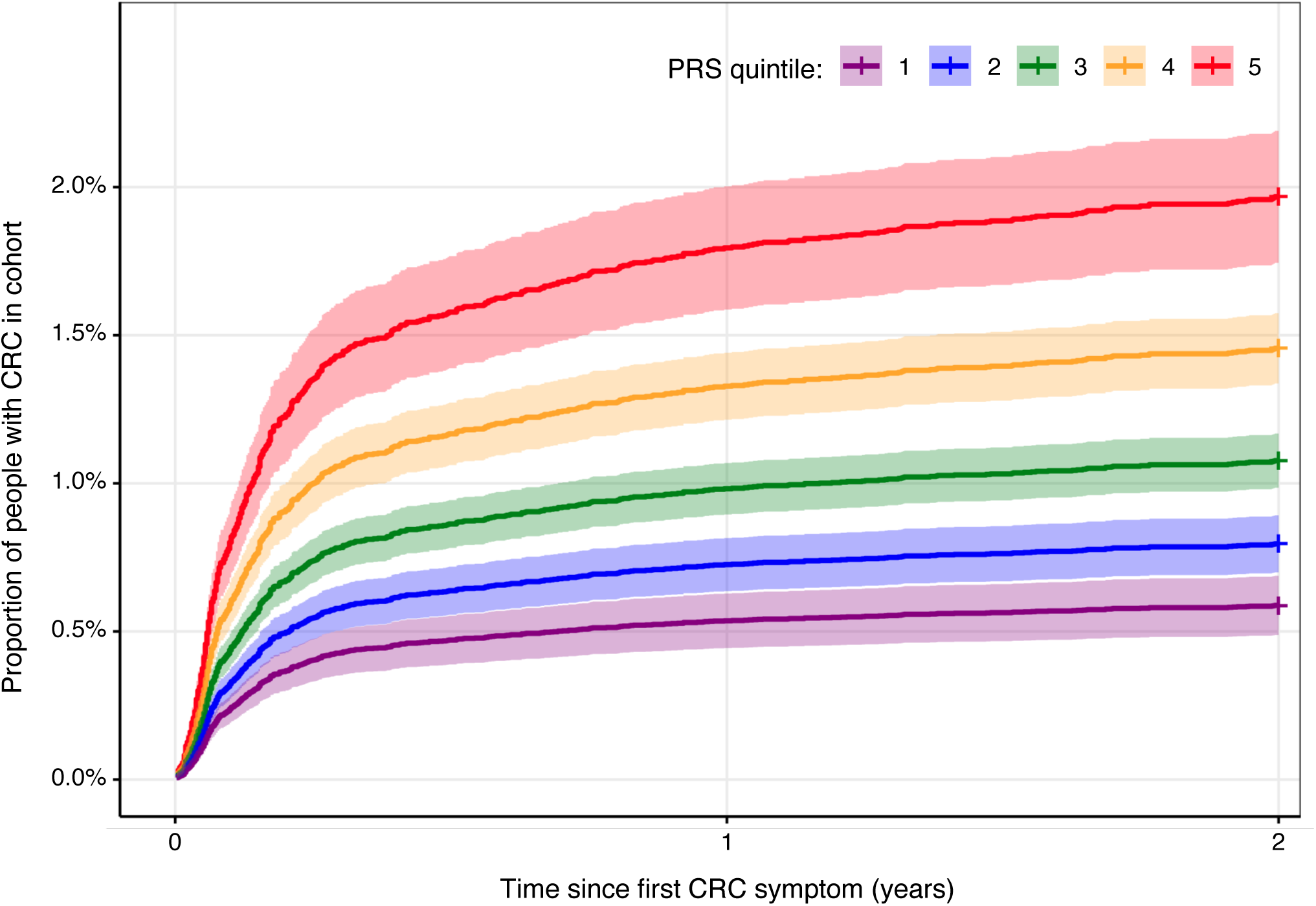
Cumulative hazard plot showing participants’ risk of CRC over two years from date of first symptom, stratified by PRS quintile. 1.95% (CI95:1.76-2.30%) of participants in the highest PRS risk quintile were diagnosed with CRC after 2 years, vs. 0.61% (CI95:0.48-0.78%) of participants in the lowest quintile. Abbreviations: CI95 = 95% confidence interval, CRC = colorectal cancer, PRS = polygenic risk score.

Predictivity of the PRS could not be assessed in non-European UKBB participants due to insufficient sample size. However, mean PRS distribution differed between ancestries in 399,454 African, Admixed American, East Asian, European, and South Asian UKBB participants (Supplementary Figure 4), being higher in Europeans. This excludes participants who were related or did not cluster into an ancestral population as described in (37), or for whom a PRS could not be calculated.

### Family history in younger participants

In male participants age 40–49, the participant self-reporting that their mother had been diagnosed with bowel cancer was the only variable which met the Bonferroni-corrected significance threshold following logistic regression (p=3.1e-6), and increased risk of CRC diagnosis within the two year period (OR=8.45, CI95=3.45–20.7). In female participants aged 40-49, there was weaker evidence (p=2.3e-02) that self-reporting a father diagnosed with bowel cancer increased risk of CRC diagnosis (OR= 3.05, CI95=1.17-7.94). However, in this subcohort, only symptom variables (abdominal pain, change in bowel habit, rectal bleeding) met the significance threshold of p<1.7e-03 (Supplementary Table 6).

## Discussion

In this study, we developed and tested a PRS and IRM in a symptomatic primary care population, for the prediction of CRC within two years of a patient first reporting a symptom. Results showed that a PRS including 207 genetic variants can predict which symptomatic patients have CRC with moderate accuracy (ROCAUC= 0.63, CI95=0.60–0.65). To our knowledge, this is the first study to use a PRS to predict short-term risk of CRC in symptomatic patients. However, numerous published PRS for long-term CRC risk have near-identical ROCAUCs of 0.629–0.631 in European populations(25–27).

1.95% of participants with PRS in the highest quintile and 0.61% in the lowest quintile were diagnosed with CRC within two years. In a 2014 study of patient perspectives, 85% of respondents chose to be investigated for CRC at a risk of 2%, indicating that this information is valuable to patients.(41)

Increased PRS was associated with higher risk of CRC diagnosis within the two years in all groups, except female participants aged 40-49. In all participants younger than 50, cases were more likely to have a parent diagnosed with bowel cancer. This may be partly explained by inherited conditions, such as Lynch syndrome, which increase risk of early-onset cancers including CRC. Jackson et al. recently highlighted the importance of family history when predicting CRC risk using genetics.(42)

The IRM developed in this study combined measures of PRS, age at symptom presentation, sex, and whether or not the patient reported symptoms of abdominal pain, rectal bleeding, or change in bowel habit. The IRM was highly predictive of CRC diagnosis within two years of first reported symptom, with a ROCAUC of 0.80 (CI95=0.78–0.81) in a cohort of European participants between the ages of 40–79. The IRM excluding the PRS has ROCAUC of 0.78 (CI95=0.76–0.80), supporting Kachuri et al.’s findings(22) that genetic variables increase risk model predictive power, although there is an overlap in confidence intervals (Supplementary Figure 5).

IRM predictivity was robust across participant groups stratified by age and sex, with ROCAUC ranging from 0.68–0.78 (mean: 0.75, SD: 0.04). Predictivity compares favourably to existing IRMs for population screening (no models predicting short-term risk of CRC diagnosis were available for comparison), with reported ROCAUC values between 0.57 and 0.78 (not including genetic biomarker tests, with ROCAUCs up to 0.88).(43)

Short-term risk prediction has potential to be immediately actionable in the clinical setting. Other than the PRS, all information included in the IRM can be collected at the primary care stage. This could inform patient triage, improving early diagnosis rates and health outcomes and reducing pressure on diagnostic secondary care services.

Recent initiatives to integrate genomic data into the UK healthcare system mean that calculation of a PRS in-clinic will become feasible for increasing numbers of patients. The NHS Genomic Medicine Service, launched in October 2018, aims to routinely offer whole-genome sequencing for genetic disorders or cancer, and is considering implementation of genomic cancer screening for asymptomatic individuals in the next five years.(44,45) The NHS Genomic Medicine Service is also aiming for data interoperability between NHS services, which would allow data collected for other health purposes to be used for risk stratification.(45) Other genomic healthcare programmes launched in the UK include Our Future Health, which will collect genomic and health data from 5 million adults,(46) and the Newborn Genomes Programme which will sequence the genomes of >100,000 newborns.(47) These initiatives demonstrate a shift in healthcare which will increase the availability of genomic data, making implementation of a PRS for risk stratification in clinic a possibility.

Neither the PRS nor IRM improve on the predictivity of FIT, which has ROCAUC 0.95 and remains the gold standard for stratification of patient risk.(48) However, patient noncompliance with FIT of 6.4–16.2% in primary care means there is a clinical need for stratification of CRC risk which doesn’t depend on FIT.(20) The IRM could be used to calculate a patient’s risk of CRC rapidly in the primary care setting, if enough information (age, sex, symptoms, and genotyping data) were available. This could reduce time to diagnosis, however, the clinical utility of this approach requires further investigation.

## Limitations

The IRM was developed and tested in European participants, due to underrepresentation of other ancestries in UKBB, which may limit the applicability of results to non-European patients. PRS distribution was higher in European individuals (Supplementary Figure 4), likely because the variants included in the PRS were discovered through a genome-wide meta-analysis of a mostly European population; these variants will therefore be more common in Europeans. Addressing this limitation will be an important focus of future work, especially considering that the IRM may be useful for CRC risk prediction in cases of FIT noncompliance, and a recent study showed FIT uptake is lower in the following ethnic groups: Asian, Black, and mixed or other.(20) This highlights the urgent need for openly available genome-wide association study data from ancestrally diverse populations, to develop accurate PRS for more individuals, and reduce health inequalities.(49) The IRM was also less predictive in patients younger than 50, likely due to lower CRC incidence in this age range. To improve health equality, further work is required to examine risk factors and markers associated with CRC in these populations.

## Conclusions

Earlier diagnosis of CRC is a priority to improve patient outcomes. Risk stratification approaches to determine which patients presenting in primary care are most likely to require diagnostic testing for CRC could increase rates of early CRC diagnosis and reduce burden on healthcare services. FIT remains the gold-standard test for prediction of CRC risk at the primary care stage. However, risk stratification methods which do not depend on FIT have the potential to improve patient outcomes in cases of patient nonadherence with the test.

The IRM developed in this study predicts, with high accuracy, which patients presenting with CRC symptoms in a primary care setting are likely to be diagnosed with CRC within the next two years. The IRM includes age, sex, and three symptoms (abdominal pain, rectal bleeding, and change in bowel habit). It also integrates a 207-variant PRS which stratifies patients with CRC incidence between 0.61% and 1.95% within two years of first symptom. The IRM was developed and tested in a mixed-sex, white European, symptomatic cohort of participants aged 40–79. Although external validation in a diverse cohort is required to test predictivity of the IRM in patients outside of this demographic, the IRM has potential to improve CRC risk prediction for the up to 16.2% of symptomatic patients missed by FIT.

## Data Availability

All participant data analysed in the study are available from UK Biobank following successful application to access the UK Biobank resource for research.
This study used 235 Read codes (NHS clinical descriptors) describing colorectal cancer symptoms. 159 of these were provided by the Diagnosis of Symptomatic Cancer Optimally (DISCO) consortium, University of Exeter and are available upon reasonable request to the authors. The remaining 76 Read codes for colorectal cancer symptoms, and 59 Read codes describing colorectal cancer diagnosis, are available online at: https://github.com/bethan-mallabar-rimmer/CRC_IRM/tree/main
All other data produced in the present work are contained in the manuscript.

## Data availability statement

This research was conducted using the UK Biobank Resource under Application Number 74981.

### 235 Read codes describing CRC symptoms

200 Read codes (filtered to 159) were provided by the Diagnosis of Symptomatic Cancer Optimally (DISCO) consortium, University of Exeter. These are available upon reasonable request.

19 additional Read codes were found by Dr. Matthew Barclay, University College London. Using the aforementioned 178 codes as input, 57 further Read codes were identified using a function built in R (see Code availability). These codes are available from: https://github.com/bethan-mallabar-rimmer/CRC_IRM/tree/main/CRC_read_codes

### 59 Read codes describing CRC

These are available from: https://github.com/bethan-mallabar-rimmer/CRC_IRM/tree/main/CRC_read_codes

## Code availability

This analysis used an R function ‘find_read_codes’ (written by Bethan Mallabar-Rimmer, University of Exeter) which takes Read 2 and/or 3 codes as input and returns similar Read codes starting with the same series of characters. Available from: https://github.com/bethan-mallabar-rimmer/CRC_IRM/tree/main/find_read_codes

A PRS was calculated using the following R functions for analysing UKBB genetic data on the RStudio Workbench implementation on DNA Nexus, written by Dr. Harry Green and Bethan Mallabar-Rimmer, University of Exeter: https://github.com/hdg204/Rdna-nexus

This study’s analysis pipeline is published at: https://github.com/bethan-mallabar-rimmer/CRC_IRM/tree/main

## Acknowledgements

The authors would like to thank Dr. Sarah Price and the Diagnosis of Symptomatic Cancer Optimally (DISCO) consortium, University of Exeter, for providing a list of Read codes for CRC symptoms which were vital to the analysis conducted in this study.

## Author Contributions

BMR, SWDM, SB and HG conceptualised and designed the study. KR, JT, ARW, MW, MB curated the dataset. BMR and HG carried out data preparation and analysis. BMR, APW and HG wrote the first draft of the manuscript. All authors reviewed the results and contributed to their interpretation and take the responsibility for the decision to submit for publication.

## Funding

This work was supported by a generous donation from the Higgins family. Study authors were supported by the National Institute for Health and Care Research Exeter Biomedical Research Centre, the University of Exeter Medical School, and Cancer Research UK.

## Ethical Approval

Data from the UK Biobank Resource was accessed under Application Number 74981. UK Biobank was approved as tissue bank resource by North West Multi-centre Research Ethics Committee. All UK Biobank participants gave written informed consent for use of their data for health research. Participants who withdrew consent during this study were excluded from analysis.

## Competing Interests

The authors declare no competing interests.

## Supplementary Figures

**Supplementary Figure 1A.**
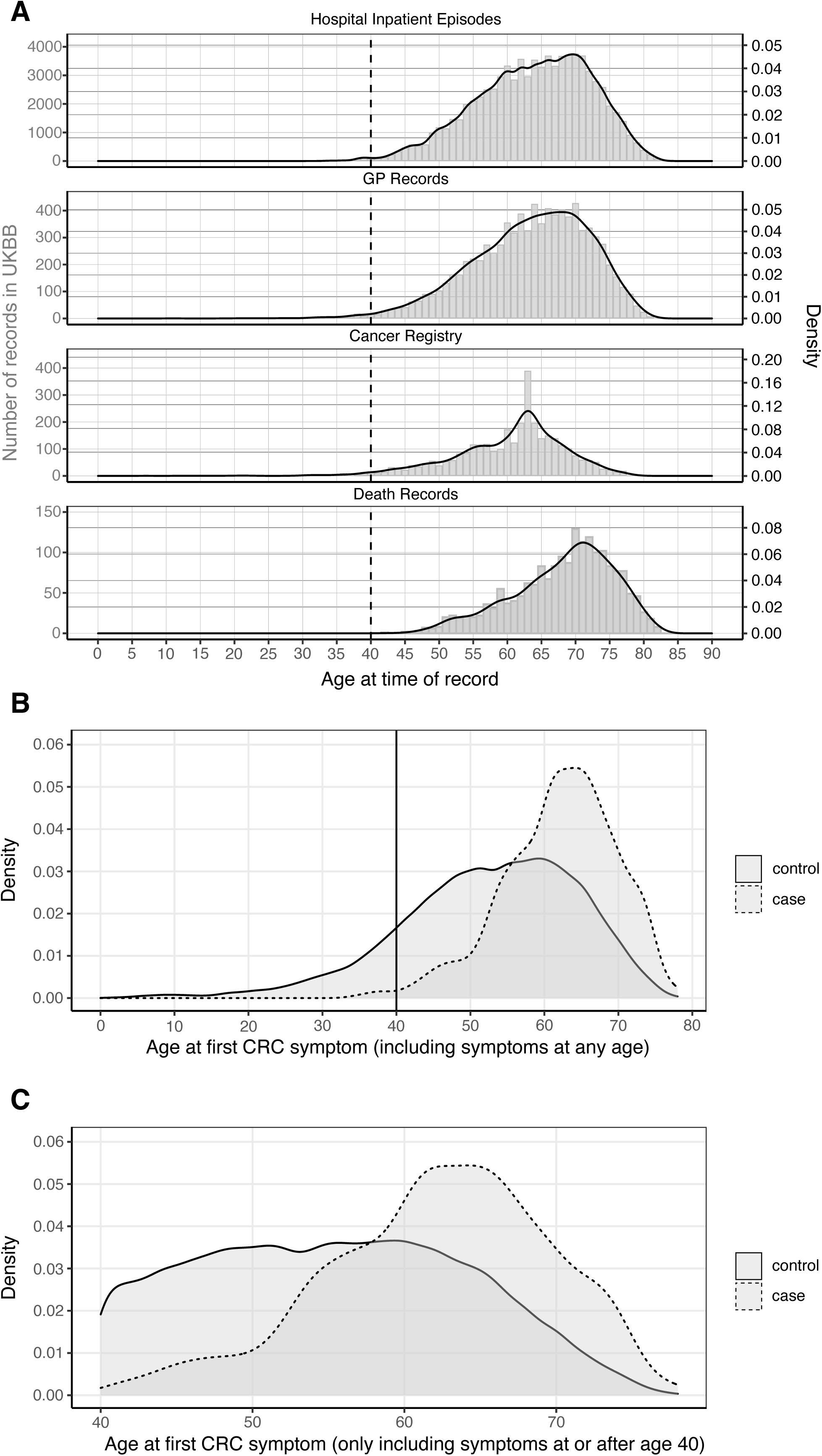
histogram and density plot of all healthcare records describing CRC in UKBB, sorted by participant age at time of record. This includes all participant records in hospital inpatient, cancer registry, and death record data labelled with ICD-10 codes C18–21, and all GP records labelled with one of 59 Read codes for CRC (see Data Availability). Duplicated health records – from the same participant, source, and date – were de-duplicated. However, if a participant had multiple diagnoses of CRC recorded, from different sources and/or dates, these records are all included in the graph. The dashed vertical line intercepts age 40, highlighting that there are very few CRC records in UKBB from individuals younger than this at the time of the record. **1b – distribution of age at first CRC symptom in cases and controls (no age threshold used when searching participants’ GP records for CRC symptoms).** Among participants whose first CRC symptom was reported at age <35, there are no cases with a diagnosis of CRC within two years of first symptom. The proportion of cases starts to increase among participants age 40+ at time of first CRC symptom, and increases sharply again among participants with first CRC symptom reported at age 50+. **1c – distribution of age at first CRC symptom in cases and controls (age threshold of 40 used when searching participants’ GP records for CRC symptoms).** This age threshold improves the proportion of cases in the cohort relative to Figure 1B, as participants with a symptom at age <40 were highly unlikely to have cancer. Abbreviations: = CRC = colorectal cancer, GP = general practice, UKBB = UK Biobank.

**Supplementary Figure 2.**
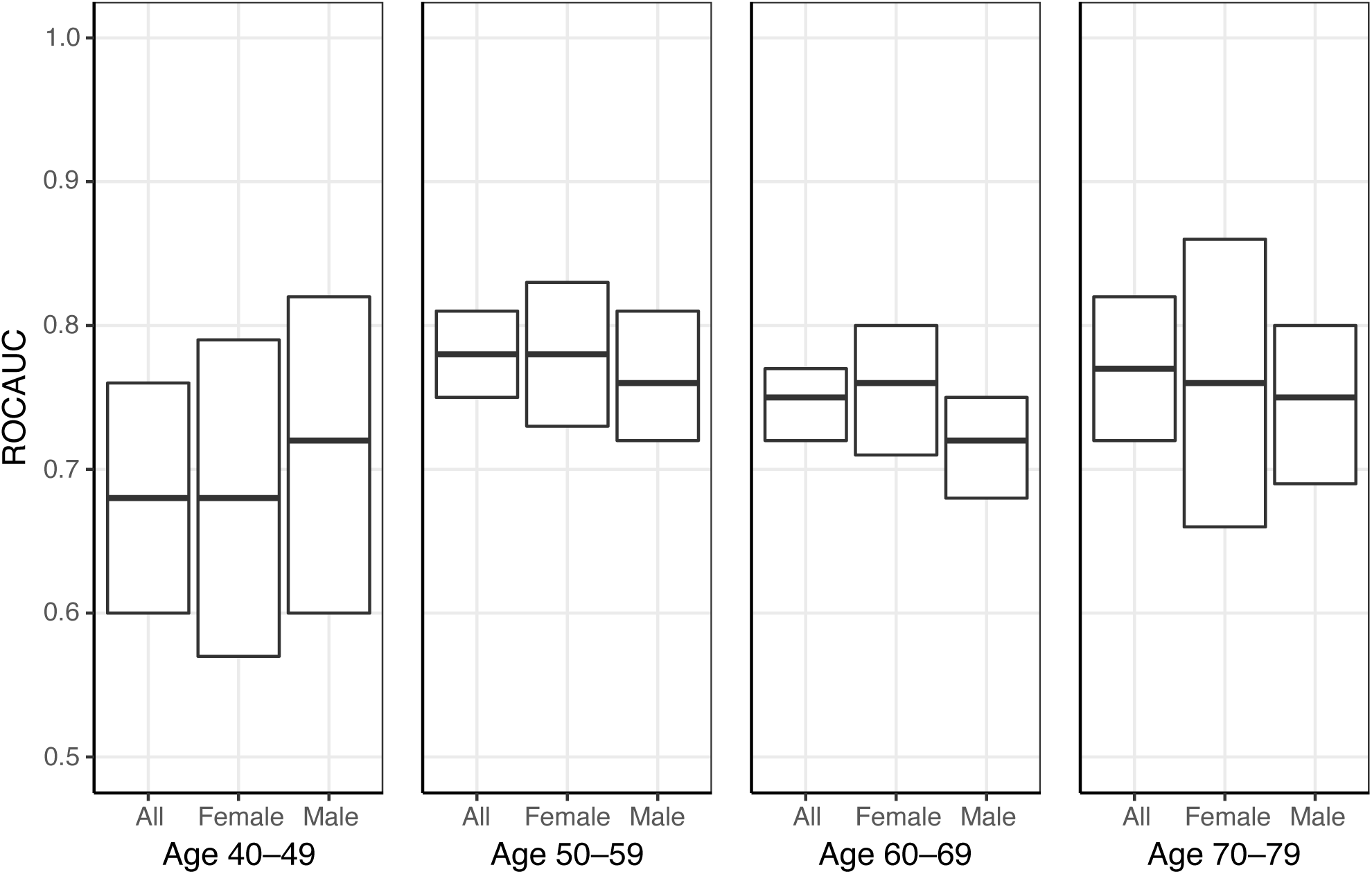
ROCAUC of the six-variable IRM across subcohorts. Middle bars show the ROCAUC of each model, upper and lower bars show the 95% confidence intervals. In cohort participants whose first CRC symptom was reported at age 40-49, the IRM shows a trend of lower predictivity and has wider confidence intervals. Abbreviations: CRC = colorectal cancer, IRM = integrated risk model, ROCAUC = receiver operating characteristic area under the curve.

**Supplementary Figure 3.**
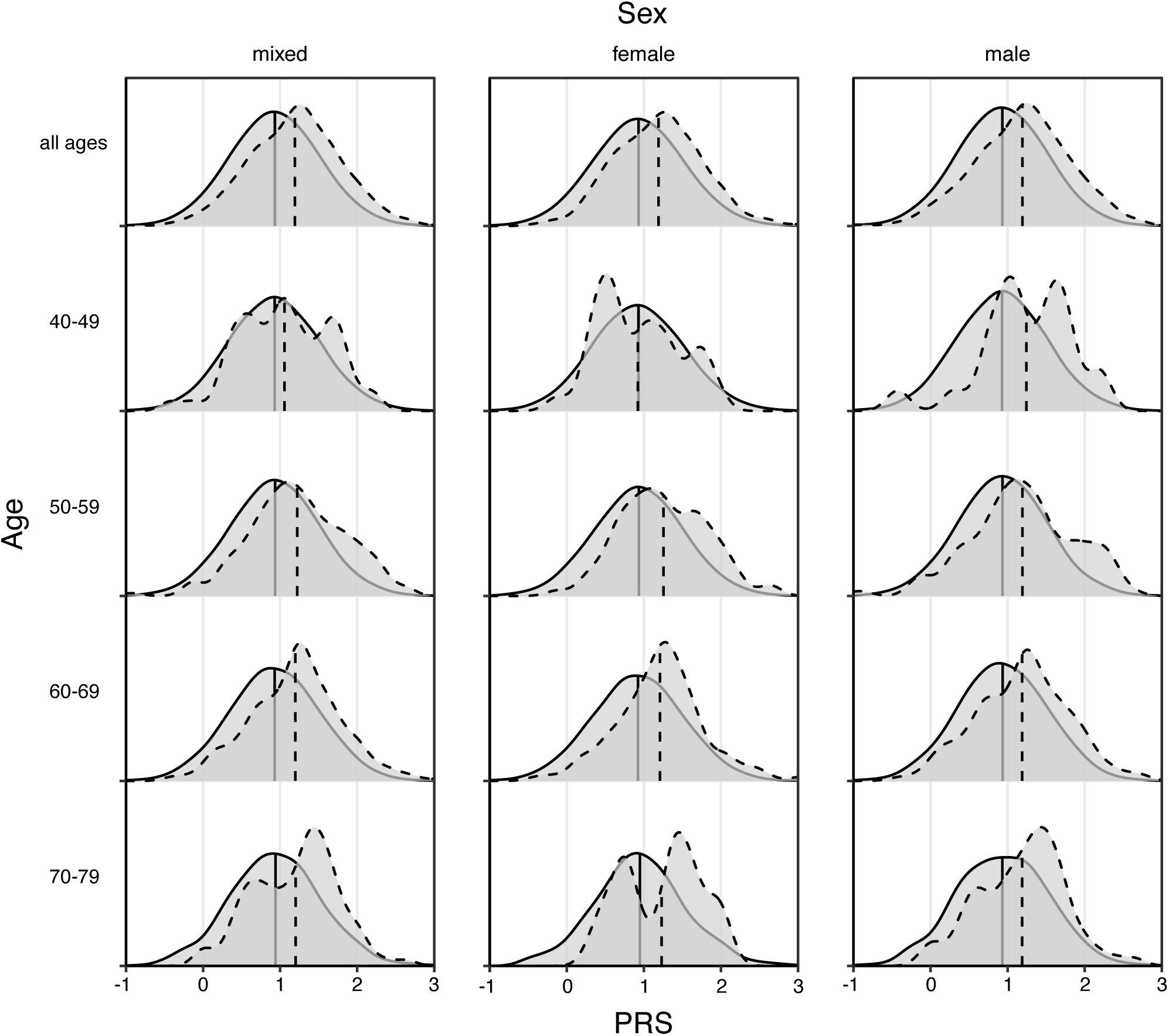
PRS distribution in cases and controls in the full cohort (top left) and subcohorts. PRS distribution is shown using a density plot, where the area under the curve represents 100% of either the case or control population in each (sub)cohort. The dashed line shows distribution in cases, the solid line in controls. Mean PRS in each cohort is shown by vertical lines. Mean PRS was significantly higher in cases than controls (Bonferroni-corrected p-value <1.7e-03), except in all three subcohorts aged 40-49 and female participants aged 70-79 (logistic regression p-values reported in Supplementary Table 6). Insignificant p-value in the latter subcohort is potentially due to low sample size (Supplementary Table 2 contains sample sizes of each cohort). Abbreviations: PRS = polygenic risk score.

**Supplementary Figure 4.**
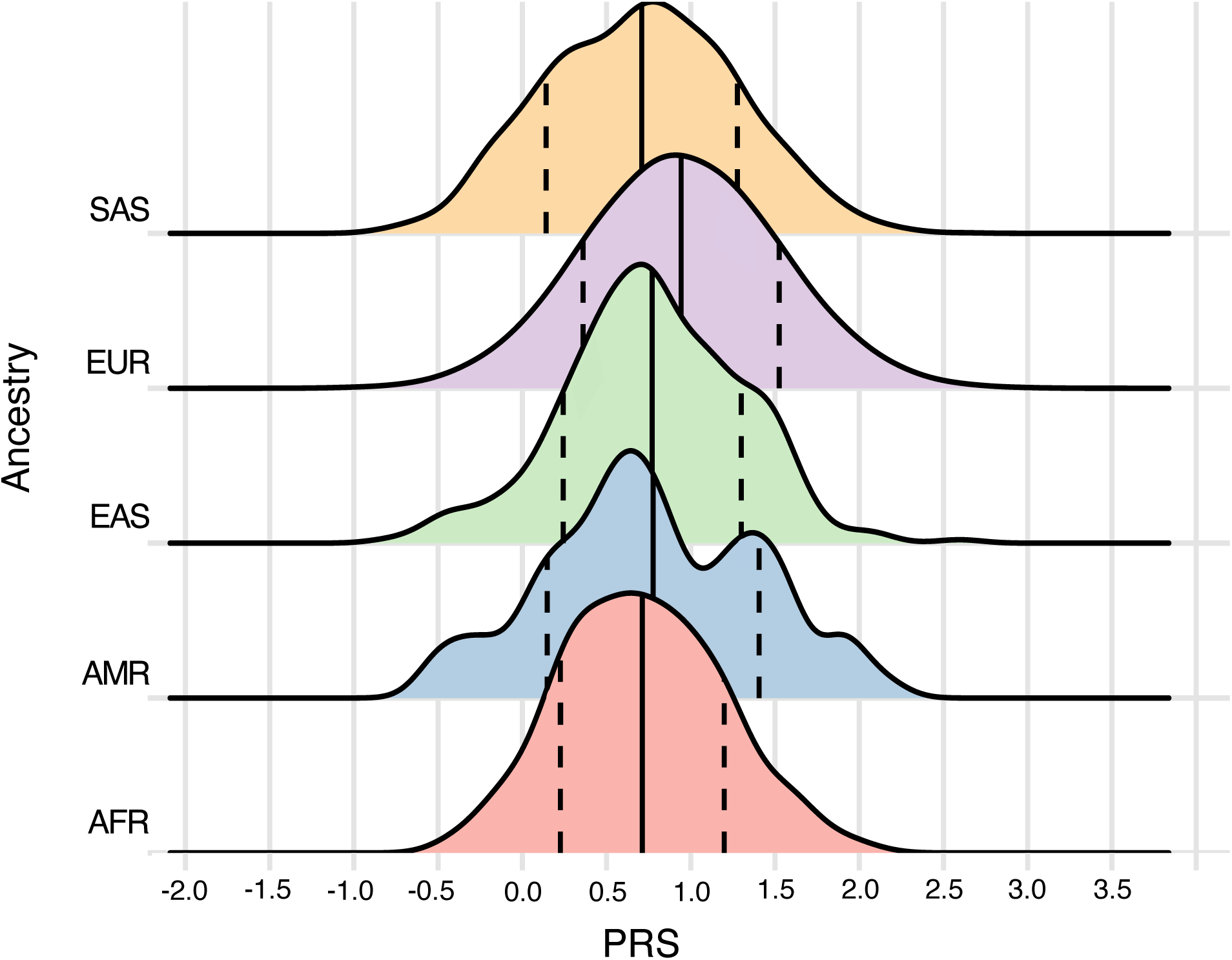
PRS differed across ancestries in 399,454 UKBB participants. This is all 502,405 UKBB participants excluding 1. individuals for whom a PRS could not be calculated, for example due to lack of genotyping data (15,207 participants excluded), 2. individuals who did not cluster into any of the five ancestral populations reported here (a further 14,747 excluded), and 3. individuals who were related to the first- or second-degree to others within the same ancestral population (a further 72,997 excluded) – method reported in (37). PRS distribution in each ancestral population is shown using a density plot, where the area under the curve represents 100% of each population. A solid vertical line shows mean PRS in each population. Dashed lines show standard deviation. A Welch’s ANOVA test (which unlike ANOVA does not assume homoscedasticity between groups) returned p <2.2e-16, showing that mean PRS differs across ancestries. Ancestral populations were compared pair-wise with Games-Howell testing (a post-hoc test for Welch’s ANOVA if group sizes are unequal, as in this cohort where non-European participants are underrepresented). This showed mean PRS in Europeans was higher than in all other groups. Mean PRS was higher in East Asian individuals than African (adjusted p = 2.90e-08) and South Asian (adjusted p = 3.58e-08) individuals; no significant difference was found compared to Admixed American participants (adjusted p = 1.21e-01). Abbreviations: AFR = African, AMR = Admixed American, EAS = East Asian, EUR = European, PRS = polygenic risk score, SAS = South Asian.

**Supplementary Figure 5.**
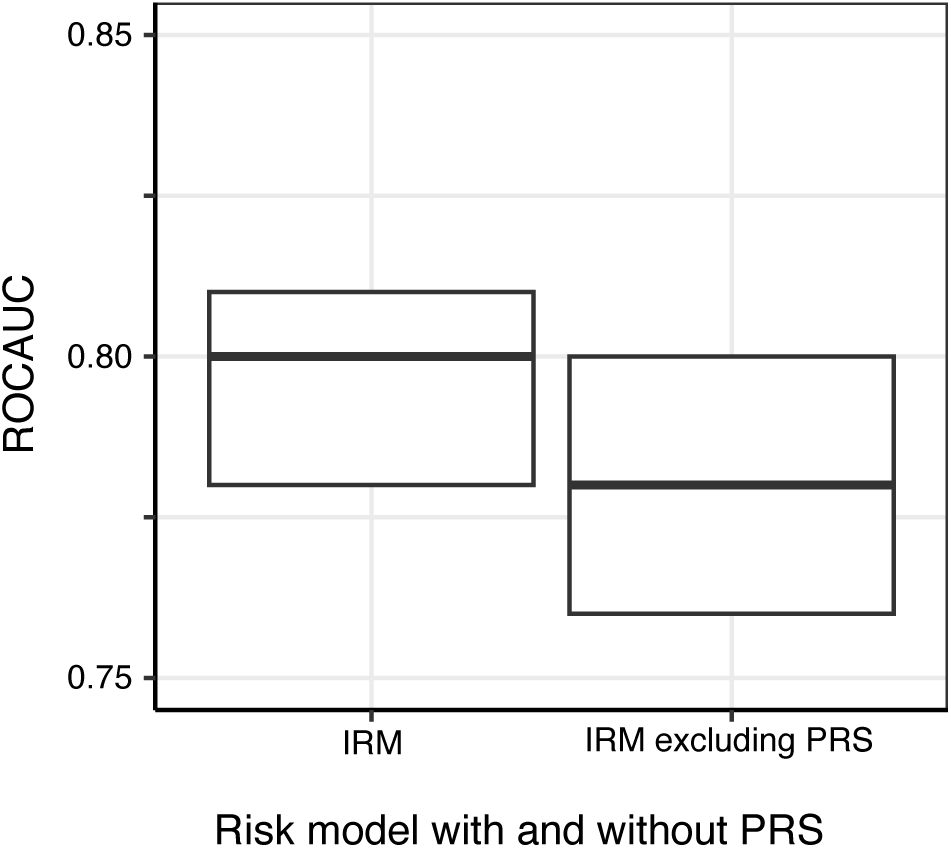
ROCAUC of the six-variable IRM developed in this study is improved through inclusion of a PRS, although confidence intervals overlap. This figure shows only the PRS and IRM developed in this study. It was based off a figure published by Kachuri et al.(23) in a systematic review of cancer risk prediction models, which showed that models including PRS performed better than ones without. The middle bar shows the ROCAUC of each model, and the upper and lower bars show the 95% confidence intervals. Abbreviations: IRM = integrated risk model, PRS = polygenic risk score, ROCAUC = receiver operating characteristic area under the curve.

## Notes

**Support received** This work was supported by a generous donation from the Higgins family. This study was supported by the National Institute for Health and Care Research Exeter Biomedical Research Centre and the University of Exeter Medical School. BMR is supported by a Cancer Research UK project grant (EDDPJT-May22\100006). SB is supported by an NIHR Advanced Fellowship (NIHR301666). The views expressed are those of the author(s) and not necessarily those of the funders named.

### Competing Interest Statement

The authors have declared no competing interest.

### Funding Statement

This work was supported by a generous donation from the Higgins family.
This study was supported by the National Institute for Health and Care Research Exeter Biomedical Research Centre and the University of Exeter Medical School. BMR is supported by a Cancer Research UK project grant (EDDPJT-May22\100006). SB is supported by an NIHR Advanced Fellowship (NIHR301666). The views expressed in the manuscript are those of the author(s) and not necessarily those of the funders named.

### Author Declarations

All human data in this study came from UK Biobank. Data from the UK Biobank resource was accessed under Application Number 74981. The North West Multi-centre Research Ethics Committee gave ethical approval for UK Biobank as a research tissue bank resource. Researchers using UK Biobank do not require separate ethical clearance and can operate under the research tissue bank resource approval.

